# Influence of milk product safety and household food hygiene on bacterial contamination of infant food in peri-urban Kenya

**DOI:** 10.1101/2020.08.24.20181222

**Authors:** Vivian Hoffmann, Sheillah Simiyu, Daniel Sewell, Kevin Tsai, Oliver Cumming, Jane Mumma, Kelly K. Baker

**Affiliations:** International Food Policy Research Institute, Washington, D.C., United States; Africa Population Health Research Center, Nairobi, Kenya; Department of Biostatistics, University of Iowa, Iowa City, Iowa, United States; Department of Occupational and Environmental Health, University of Iowa, Iowa City, Iowa, United States; Department of Disease Control, London School of Hygiene and Tropical Medicine, London, United Kingdom; Department of Community Nutrition, Great Lakes University of Kisumu, Kisumu, Kenya

**Author notes:** Corresponding author: Kelly K. Baker, University of Iowa College of Public Health, 145 S. Riverside Dr., S316, Iowa City, IA 52246; (001).

## Abstract

**Background:** Previous work in peri-urban communities within Kisumu County, Kenya, has shown that milk is a common weaning food and often contains enteric pathogens. Little is known about how milk and milk-based foods fed to infants become contaminated in peri-urban communities.

**Objective:** To compare the bacterial indicator and enteric pathogen detection levels in unpackaged, fresh pasteurized, and ultra-high temperature (UHT) treated milk at purchase, and assess its association with contamination of food prepared with this milk and fed to infants.

**Methods:** Paired samples of milk at point of sale and infant food prepared with this milk were obtained from 188 households enrolled as controls in the Safe Start trial. Samples were cultured to isolate *Salmonella enterica, Shigella sonnei, Klebsiella aerogenes, Proteus* spp., and *E. Cole* including Enterohemorrhagic (EHEC) *E. coli* 0157, with pathogens validated by PCR. Detection of these bacteria was compared across milk types, and between milk at purchase vs. point of infant consumption.

**Results:** Unpackaged milk was most contaminated at point of purchase, but bacterial contamination was also present in pasteurized and UHT milk. Presence of bacteria in milk at purchase predicted presence of the same bacteria type in infant food. Contamination detection frequency and concentration level for bacterial indicators generally increased between point of purchase and consumption among users of UHT and fresh pasteurized milk, but decreased among those using unpackaged milk. Detection of the four fecal bacteria in infant food was not significantly related to the type of milk used.

**Conclusion:** Kenyan infants are exposed to foodborne bacteria, including enteric pathogens, in milk-based foods. Both pre-market contamination and post-purchase handling influence the likelihood of milk given to infants being contaminated. Improvements in the safety of raw and pasteurized milk, and caregiver education on safe preparation and storage, are needed to reduce infant exposure to contaminated food.

**What is already known?:** Globally, food is an important cause of diarrheal outbreaks although evidence on the importance of food as a source of child exposure to diarrheal pathogen, and the source of those pathogens, in low income nations is scarce. While household hygiene influences infant food safety, consumption of pre-packaged foods, like pasteurized milk, have grown rapidly in recent decades, especially in urban populations, highlighting possible roles for both food systems and households in foodborne exposure of children.

**What are the new findings?:** Kenyan infants are frequently exposed to foodborne bacteria, including enteric pathogens, in milk-based supplemental foods. Pre-packaged pasteurized milk products are safer at point of purchase than unpackaged milk, although offered no protection for infants against bacterial exposure at point of consumption. Both purchased milk products and post-purchase household food handling behaviors influenced the likelihood of infant exposure to contaminated infant food.

**What do the new findings imply?:** Joint interventions targeting improvements in the safety of raw and pasteurized milk production, as well as caregiver education on safe preparation and storage of food, are needed for preventing foodborne exposure of peri-urban Kenyan infants.

## INTRODUCTION

Morbidity and mortality among children under 5 years of age accounts for 40% of the global burden of foodborne disease ^1^. Bacterial contamination in weaning foods is typically higher in concentration than drinking water ^2 3^, and there is evidence that weaning foods are more contaminated than foods consumed by adults ^4^. Milk may be particularly high risk to infants because it frequently contains a higher number of fecal conforms than many other foods ^2 5 6^. Recent research in Kenya, where cow’s milk is an important weaning food, shows that milk stored in the household for feeding to young children is more likely to be contaminated with an enteric pathogen than other weaning foods ^7^. An important question for addressing the problem of unsafe weaning foods is the source of this contamination: Does milk already contain pathogens when it is brought into the household, or do these enter under unhygienic household food preparation, treatment, and storage conditions?

Previous quantitative risk assessments of disease transmission through milk in Kenya have focused on informally marketed raw milk. These have used self-reported boiling behavior to estimate the proportion of households who boil milk prior to consumption and have assumed that boiling is 100% effective against the pathogens studied ^8, 9^. As the vast majority of households typically report boiling milk purchased from the informal market prior to consumption, previously estimated risks of exposure to foodborne illness through milk consumption have been small. However, these previous studies do not consider several important factors. First, consumption of pasteurized milk is high among Kenya’s rapidly growing urban population. Nationally representative data collected in 2015 show that 66% of households in Nairobi, Kenya’s largest city, and 60% of those in peri-urban Kisumu, the site of the present study, had consumed processed milk over the past 7 days ^10^. An earlier study found that while consumption of pasteurized milk in Nairobi increased with income, even households in the lowest income quintile were as likely to consume pasteurized as raw milk ^11^. Due to the perception that it is ready to drink, packaged milk may not be boiled, even by those who typically boil unpackaged, informally marketed milk.

Second, processed milk may be safer than raw milk, but still contain pathogens. Some pathogens, such as heat-resistant spores of *C. botulinum* or *B. cereus*, can survive pasteurization (heated to 71-74 °C for 15-40 seconds) ^12^. Ultra-high temperature (UHT) “long life” treatment of milk (135-140°C for 6-10 seconds) more efficiently destroys vegetative pathogens and heat-resistant spore forming pathogens, but is still not full sterilization. Yet, the quality of UHT milk allows it to be stored at room temperature for longer periods of time, which has reduced the cost of packaged milk production and transport, expanded the UHT distribution market into informal settlements and rural areas, and contributed to the rise in its global consumption. Further, processed milk may be contaminated with pathogens after treatment. This may occur if processing equipment is not properly cleaned, if packaging materials are contaminated, or if the very low levels of pathogens remaining in milk post-pasteurization are able to multiply due to failures in the cold chain. *Listeria monocytogenes* from biofilms on processing equipment have caused foodborne outbreaks in several settings ^13^.

Third, survey respondents may misreport boiling behavior to researchers due to a desire to be seen to be doing the “right thing”. Finally, even if households are boiling milk prior to consumption, recontamination of the boiled milk may occur through utensils and hands which have come into contact with un-boiled milk or household surfaces or airborne dust. If contamination is bacterial, then contamination levels can rise over time with bacterial replication if milk is stored at room temperature. As a single batch of prepared infant food is often consumed over multiple feeding events, spanning several hours, the potential for bacterial replication during storage is high.

The aim of this study is to assess food safety risks associated with different types of purchased milk given to infants at eight months of age in Kenya, where diarrheal disease mortality is high ^14^. Specifically, we assess the contributions of microbial contamination at time of purchase, and contamination introduced during use within the household, to overall risk of contamination of milk-based infant food.

## METHODS

### Study Design

The Market to Mouth study builds on a cluster-randomized randomized controlled trial, Safe Start, based in the Nyalenda A and Nyalenda B wards of peri-urban Kisumu. These densely populated wards are characterized by lack of improved sanitation facilities, use of county-provided water points, poor housing, and high rates of poverty. The Safe Start study evaluates the effect of a food hygiene intervention targeting early childhood exposure to enteric pathogens through contaminated food (ClinicalTrials.gov ID: NCT03468114) ^15^. The protocol includes a midline visit at 8 months of age to observe the caregiver prepare food and feed the child, and to take a food sample for microbial testing. The Market to Mouth study uses food sample data collected at this midline visit, and from the market vendors where caregivers procured milk fed to the infants tracked by Safe Start, to examine pathogen transmission patterns. In this manuscript, we analyze 396 milk samples purchased directly from vendors patronized by Safe Start households, and matched infant food samples from the 188 households among these assigned to the Safe Start control group who reflect mothers in this population in the absence of a food safety intervention.

### Human Subjects Research

Approval for the collection of infant food samples was obtained from Great Lakes University of Kisumu (Ref: GREC/010/248/2016), London School of Hygiene and Tropical Medicine (Ref: 14695), and the University of Iowa (Ref: 00000099). An information consent form was read to caregivers in their preferred language. If caregivers consented to participate in the study, they signed consent forms in the presence of a community witness and were given copies of the informed consent form for their records. Participants were allowed to withdraw at any time.

### Patient and Public Involvement

Formative research on caregiver experiences and infant food safety ^7 16^, and the challenges for Community Health Volunteers (CHVs) in delivery of health care information ^17^ informed study design. The design of the Safe Start intervention was optimized through an interactive pilot study ^18^ and CHVs were involved in the implementation of the study ^15^. Community knowledge dissemination meetings were convened after the study to discuss results with the community.

### Data Collection and Food Sampling

When scheduling the Safe Start midline visit, the research team inquired about what the caregiver intended to prepare for the child to eat the following day. If the caregiver planned to feed the child milk or food prepared with milk, the team arranged with the caregiver for a study enumerator to meet and travel with the caregiver to procure the milk, whether this was done the same evening, the following morning, or just before the food was prepared. Only caregivers who fed their infants milk or food made with milk, for example milk tea or milk porridge, are included in the present study (54% of 733 Safe Start caregivers). The enumerator accompanying the caregiver during milk purchase recorded the type of milk (unpackaged, fresh packaged, UHT, or infant formula), price paid, volume obtained, and whether the milk was refrigerated. The enumerator then purchased additional milk of the same type from the vendor for laboratory analysis.

During the Safe Start midline visit, which typically occurred in the morning, the Safe Start research team observed the caregiver’s food preparation and infant feeding practices and collected a food sample that was transported to the laboratory for analysis. In addition to the morning visit, the Safe Start team scheduled a time in the afternoon, per convenience of the caregiver, to collect another sample of the infant food that had been prepared in the morning if it was still being used by that time. The afternoon visit was scheduled as late as possible, to capture the full influence of household storage practices on the microbial quality of infant food. The afternoon food sample was used unless it was completely consumed, in which case the morning sample was processed for all assays (n=25).

Caregivers were asked to provide a spoonful of solids (approximately 5 grams) or ~ 2 fluid ounces of this food to minimize unnecessary oversampling of food that would otherwise be fed to the infant. Both vendor and household samples were barcode labeled to match key identifiers of the household ID. All samples were transported to the laboratory on ice packs in a cooler within four hours of collection.

### Laboratory methods

Samples were processed within 2 hours of receipt by Alkaline Phosphatase (ALP) assays to assess pasteurization status, by bacterial pre-enrichment and culture assays for select foodborne pathogens and microbial indicators, and direct DNA and RNA extraction for quantitative molecular analysis for a broad array of enteric pathogens and human and bovine microbial source tracking markers.

#### Pasteurization assay

ALP assays (Charm Sciences, Inc., Lawrence, MA) were used to determine whether milk or foods containing milk had been sufficiently boiled at either high enough temperatures or long enough time periods to achieve pasteurization conditions for a subset of 38 vendor samples.

#### Analysis of Enteric Pathogen Contamination

*Culture Assays:* A 2 ml sample of liquid food or 300 mg of solid food was mixed with 2 ml of a Peptone enrichment media and incubated at 4°C for 24 hours to recover injured but viable bacteria, without promoting significant increases in replication to allow quantification of contamination. The next day, samples were incubated at 42°C for one hour to trigger bacterial replication metabolism. Microbial presence and concentration in food samples were determined using culture-based isolation and phenotyping of a subset of bacteria commonly found in the digestive tracts and feces of humans and cattle, and common foodborne pathogens: *Salmonella spp*. (including 5. *enterica*), *Shigella sonnei* (5. *sonnei*), *Enterobacter aerogenes* (otherwise known as *Klebsiella aerogenes*), general *E. coli* and pathogenic EHEC 0157, and *Proteus* spp. After pre-enrichment, 1 ml, 100 μl, and 10 μl serial dilutions volumes of sample were vacuum filtered through 0.45 μM membrane filters (Millipore, MA, USA) and cultured on the selective and differential chromogenic medium, *E. coli* 0157: H7 MUG agar (Sigma-Aldrich, #44782, St. Louis, MO, USA) for 24 hours at 35-37°C. Presumptive bacterial pathogen presence was determined by counting individual colony forming units (cfu) for each phenotype, according to the manufacturer’s protocol. One negative environmental control was performed on each day of processing. Validated bacterial strains of each target organism were used as positive controls to verify performance of each batch of pre-enrichment media or media plates, and to confirm pathogen identity during PCR.

#### Validation of Pathogen phenotype

Five colonies of each phenotype were picked into 100 ul of molecular grade water and boiled for 5 minutes to destroy bacteria cell structure and release its DNA. DNA were frozen at −20C until a PCR test could be performed to verify strain type. A qualitative polymerase chain reaction (PCR) assay was used to determine whether the *Salmonella* spp. phenotypes were pathogenic 5. *enterica (ttr* gene), and whether EHEC 0157 *(rdbE)* and the *Shigella sonnei (virG* and *ipaH)* phenotypes possessed pathogenic virulence genes. Failure to detect these genes meant samples were classified as negative forS. *enterica*, EHEC 0157, and 5. *sonnei*, respectively.

### Statistical Analysis

Statistical analysis was performed using Stata version 16.0 (StataCorp, 2019). We use Fisher’s exact test to compare the probability of microbial detection across milk types (unpackaged, fresh packaged, and UHT) and across brands at purchase, and across infant foods prepared with these types of milk. A likelihood ratio test based on a negative binomial regression model with milk type indicators is used to compare microbial diversity across milk types. As a robustness test of the comparisons across milk types, we estimate marginal effects of vendor milk type on microbial presence and diversity, controlling for modifiers (refrigeration status for vendor milk, and food type for infant food samples), using logistic and negative binomial regressions respectively.

For tests of paired data (vendor samples and paired infant food samples), we use McNemar’s exact test for binary indicators of microbial presence, and a negative binomial generalized linear model (GLM) with correlated standard errors within paired samples for microbial diversity.

To analyze the influence of microbial presence in vendor milk on that of infant food, we calculate the odds ratio for detection of each organism in infant food based on whether the same organism was found in the milk used to prepare it. To characterize changes in microbial concentration between vendor milk and infant food samples, we first exclude observations for which a given organisms is detected in neither the vendor milk nor the infant food sample, or for which microbial concentration at the maximum limit of analysis is observed in both samples. We then assess whether the proportion of paired samples in which microbial concentration increased vs. decreased between purchase and child feeding is influenced by vendor milk type using Fisher’s exact test.

The East African Community (EAC) milk standard sets a maximum acceptable *E. aerogenes* coliform bacteria plate count for pasteurized (10 cfu/ml) and raw (50,000/ml) milk ^19 20^. The total coliform plate count could be higher than the *E. aerogenes* plate count, so this provides a lower bound of true noncompliance with this element of the EAC standard. In addition to the proportion of samples containing any *E. aerogenes*, we report the proportion of samples that exceeded the coliform standard based on this organism alone.

## RESULTS

### Sources of milk and types of milk used for infant food

Of the 396 samples of milk procured for feeding to study infants, 90.4% were purchased from small shops known as dukas, 5.3% from milk bars, 0.5% from roadside vendors, 1.0% from study households’ own cows, 1.8% from neighbors’ cows, and 0.5% from larger shops. The most common type of milk purchased for infant feeding was long-life UHT milk, at 70% of all samples, followed by packaged fresh (pasteurized) milk at 21%, and unpackaged cow’s milk at 8.6%. Infant formula was used by a single caregiver. Almost all pasteurized and UHT milk and the single observation of baby formula was purchased from dukas. Most of the unpackaged milk was purchased from milk bars (21 cases), while two caregivers purchased such milk from a roadside vendor, and the remainder (11 cases) obtained unpackaged milk from a neighbor or the household’s own cow.

### Vendor practices and contamination in vended milk

Among milk types that require refrigeration (non-UHT), 77% of samples purchased from dukas were refrigerated and 76% of those purchased from milk bars were refrigerated. The two roadside vendors and neighbors from whom milk was purchased informally did not refrigerate the milk offered for sale. The single larger shop in the sample from which fresh milk was purchased also failed to refrigerate. The storage container in which vendors kept milk was observed for 32 of the 34 samples of unpackaged milk. Of these, most vendors (62.5%) stored milk in wide-mouthed containers with lids, 16.3% kept it in containers ready for sale, and 22% stored milk in wide-mouthed containers without lids, which could allow flies to enter. ALP assays of 37 packaged milk samples indicated that this milk was heated to pasteurization temperatures sufficient to kill most bacteria.

At least one type of bacteria was cultured from 21.5 % (mean 0.41 types of species/sample) of fluid milk (Table 1). No bacteria were cultured from the one infant formula. Probability and diversity of bacterial contamination were significantly associated with milk type, with the lowest risk of contamination in UHT milk and the highest in unpackaged milk.

*E. aerogenes* was the most commonly identified organism (12% of samples), and 5.6% of samples exceeded the EAC coliform standard. Fresh packed (17%), UHT (5.4%), and unpackaged milk (56%) had significant rates of contamination with *E. aerogenes*. Fresh pasteurized milk was more likely to exceed the EAC coliform standard than UHT milk but less likely than unpackaged milk, noting though that unpackaged milk has a higher EAC standard compliance thresholds.

*Salmonella* spp. were cultured in 7.3% of fresh milk samples, 5.3% of which were human pathogen 5. *enterica. S. enterica* was significantly more common in unpackaged milk (41%, n=34) than either packaged fresh (2.4%, n=83) or UHT milk (1.8%, n=278). Among the 21 5. *enterica* positive samples, the concentration of 62% was below 100 cfu/mL, 9.5% between 100 and 1000 cfu/mL, 21% between 1000 and 10,000 cfu/mL, and 9.5% above 100,000 cfu/mL.

All *Shigella sonnei* were *ipaH* positive, suggesting human pathogenicity. 5. *sonnei* contamination was also most common in unpackaged milk at 59% of samples, compared to 3.6% of fresh pasteurized milk and 0.7% of UHT milk. Among the 25 5. *sonnei* positive samples, the concentration of 40% were below 100 cfu/mL, 36% between 100 and 1000 cfu/mL, 4% between 1000 and 10,000 cfu/mL, and 20% above 100,000 cfu/mL^1^.

One packaged fresh milk sample was positive for *Proteus* spp. (26 cfu/ml), none were positive for general *E. coli*, and 15% were positive for the EHEC 0157 phenotype. All presumptive EHEC 0157 colonies were negative for the *rdbE* gene and could not be validated as a pathogen health hazard. Due to the lack of variability in general *E. coli* and *Proteus* spp. these outcomes are omitted from the subsequent analysis aside from their inclusion among the organisms used to assess presence of any bacteria and bacterial diversity in infant food (Table 3).

Distributional requirements for statistical tests of differences in microbial concentration across milk types were not met. However, for most of the organisms analyzed the mean of logi_0_-transformed cfu/ml is generally highest in unpackaged milk (Figure S1). Adjusting the comparisons of microbial detection and diversity for vendor refrigeration practices and brand through a multivariate logistic regression model did not significantly affect the rate of microbial detection or microbial diversity (Table S1), and differences across milk type are similar to those shown in Table 1.

Sixteen brands of milk were purchased by caregivers and rates of noncompliance with the EAC coliform standard were statistically significant different across common brands (Table 2, p<0.001). Further brand analysis are reported in Supplemental Material.

**Table 1.**
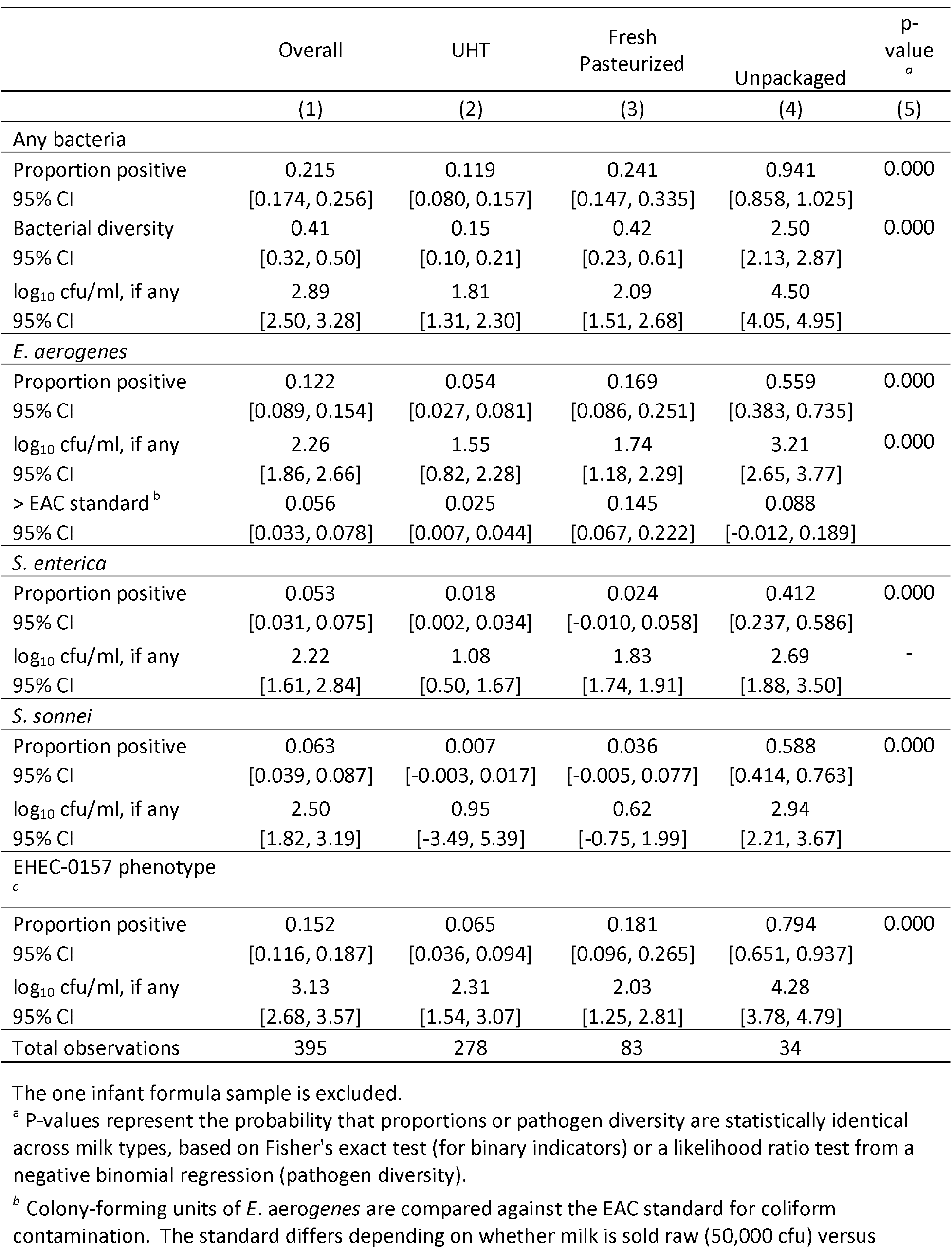
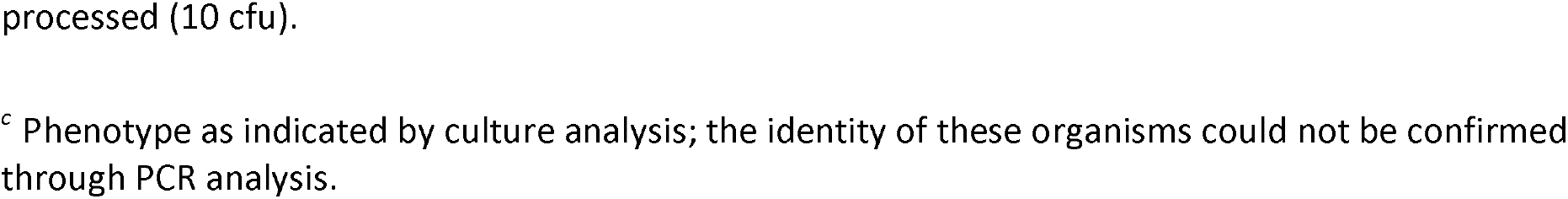
Microbial detection rates, diversity, and concentrations in 395 fluid vendor milk samples at purchase, by milk treatment type.

|  | Overall | UHT | Fresh<br>Pasteurized | Unpackaged | p-value<br><sup>a</sup> |
| --- | --- | --- | --- | --- | --- |
|  | (1) | (2) | (3) | (4) | (5) |
| *Any bacteria* |  |  |  |  |  |
| Proportion positive | 0.215 | 0.119 | 0.241 | 0.941 | 0.000 |
| 95% CI | [0.174, 0.256] | [0.080, 0.157] | [0.147, 0.335] | [0.858, 1.025] |  |
| Bacterial diversity | 0.41 | 0.15 | 0.42 | 2.50 | 0.000 |
| 95% CI | [0.32, 0.50] | [0.10, 0.21] | [0.23, 0.61] | [2.13, 2.87] |  |
| log<sub>10</sub> cfu/ml, if any | 2.89 | 1.81 | 2.09 | 4.50 |  |
| 95% CI | [2.50, 3.28] | [1.31, 2.30] | [1.51, 2.68] | [4.05, 4.95] |  |
| *E. aerogenes* |  |  |  |  |  |
| Proportion positive | 0.122 | 0.054 | 0.169 | 0.559 | 0.000 |
| 95% CI | [0.089, 0.154] | [0.027, 0.081] | [0.086, 0.251] | [0.383, 0.735] |  |
| log<sub>10</sub> cfu/ml, if any | 2.26 | 1.55 | 1.74 | 3.21 | 0.000 |
| 95% CI | [1.86, 2.66] | [0.82, 2.28] | [1.18, 2.29] | [2.65, 3.77] |  |
| > EAC standard<sup>b</sup> | 0.056 | 0.025 | 0.145 | 0.088 |  |
| 95% CI | [0.033, 0.078] | [0.007, 0.044] | [0.067, 0.222] | [-0.012, 0.189] |  |
| *S. enterica* |  |  |  |  |  |
| Proportion positive | 0.053 | 0.018 | 0.024 | 0.412 | 0.000 |
| 95% CI | [0.031, 0.075] | [0.002, 0.034] | [-0.010, 0.058] | [0.237, 0.586] |  |
| log<sub>10</sub> cfu/ml, if any | 2.22 | 1.08 | 1.83 | 2.69 | - |
| 95% CI | [1.61, 2.84] | [0.50, 1.67] | [1.74, 1.91] | [1.88, 3.50] |  |
| *S. sonnei* |  |  |  |  |  |
| Proportion positive | 0.063 | 0.007 | 0.036 | 0.588 | 0.000 |
| 95% CI | [0.039, 0.087] | [-0.003, 0.017] | [-0.005, 0.077] | [0.414, 0.763] |  |
| log<sub>10</sub> cfu/ml, if any | 2.50 | 0.95 | 0.62 | 2.94 |  |
| 95% CI | [1.82, 3.19] | [-3.49, 5.39] | [-0.75, 1.99] | [2.21, 3.67] |  |
| EHEC-0157 phenotype<sup>c</sup> |  |  |  |  |  |
| Proportion positive | 0.152 | 0.065 | 0.181 | 0.794 | 0.000 |
| 95% CI | [0.116, 0.187] | [0.036, 0.094] | [0.096, 0.265] | [0.651, 0.937] |  |
| log<sub>10</sub> cfu/ml, if any | 3.13 | 2.31 | 2.03 | 4.28 |  |
| 95% CI | [2.68, 3.57] | [1.54, 3.07] | [1.25, 2.81] | [3.78, 4.79] |  |
| Total observations | 395 | 278 | 83 | 34 |  |

The one infant formula sample is excluded.
<sup>a</sup> P-values represent the probability that proportions or pathogen diversity are statistically identical across milk types, based on Fisher's exact test (for binary indicators) or a likelihood ratio test from a negative binomial regression (pathogen diversity).
<sup>b</sup> Colony-forming units of *E. aerogenes* are compared against the EAC standard for coliform contamination. The standard differs depending on whether milk is sold raw (50,000 cfu) versus
processed (10 cfu).
<sup>c</sup> Phenotype as indicated by culture analysis; the identity of these organisms could not be confirmed through PCR analysis.

**Table 2.**
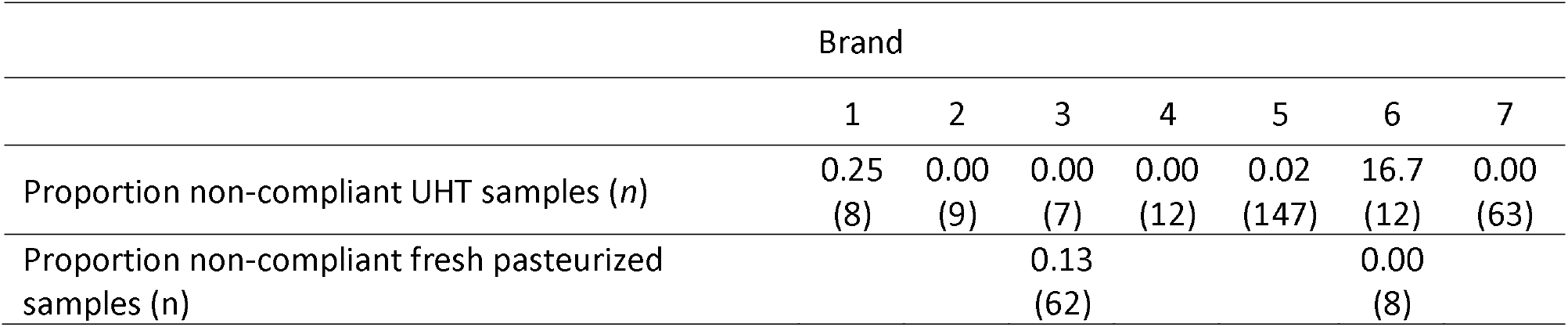
Non-compliance by brand and milk type, EAC coliform standard (based on *E. aerogenes* cfu/ml)

|  | Brand |  |  |  |  |  |  |
| --- | --- | --- | --- | --- | --- | --- | --- |
|  | 1 | 2 | 3 | 4 | 5 | 6 | 7 |
| Proportion non-compliant UHT samples (*n*) | 0.25<br>(8) | 0.00<br>(9) | 0.00<br>(7) | 0.00<br>(12) | 0.02<br>(147) | 16.7<br>(12) | 0.00<br>(63) |
| Proportion non-compliant fresh pasteurized samples (*n*) |  |  | 0.13<br>(62) |  |  | 0.00<br>(8) |  |

### Contamination in infant food

After household handling and storage of infant food made with milk, contamination rates across milk types did not differ at the statistical threshold of p<0.05 (Table 3). Pooling observations across all types of milk, bacteria were cultured from 60% of infant food samples, with a mean of 1.3 bacterial species phenotypes per sample, and a total count of logio 3.2 cfu/ml. Both the proportion of infant foods in which any bacteria were detected (McNemar’s exact test), and average bacterial diversity (negative binomial GLM allowing correlation within paired samples), are significantly higher than in vended milk overall (p<0.001). *E. aerogenes, S. enterica, S. sonnet* and the EHEC 0157 phenotype were detected in 41% (37% above guidelines), 7%, 21%, and 48% of infant foods, respectively. Among these, the difference in the proportion of samples between vended milk and infant food prepared with that milk was significant for all bacteria at p<0.01, except for 5. *enterica*.

**Table 3.**
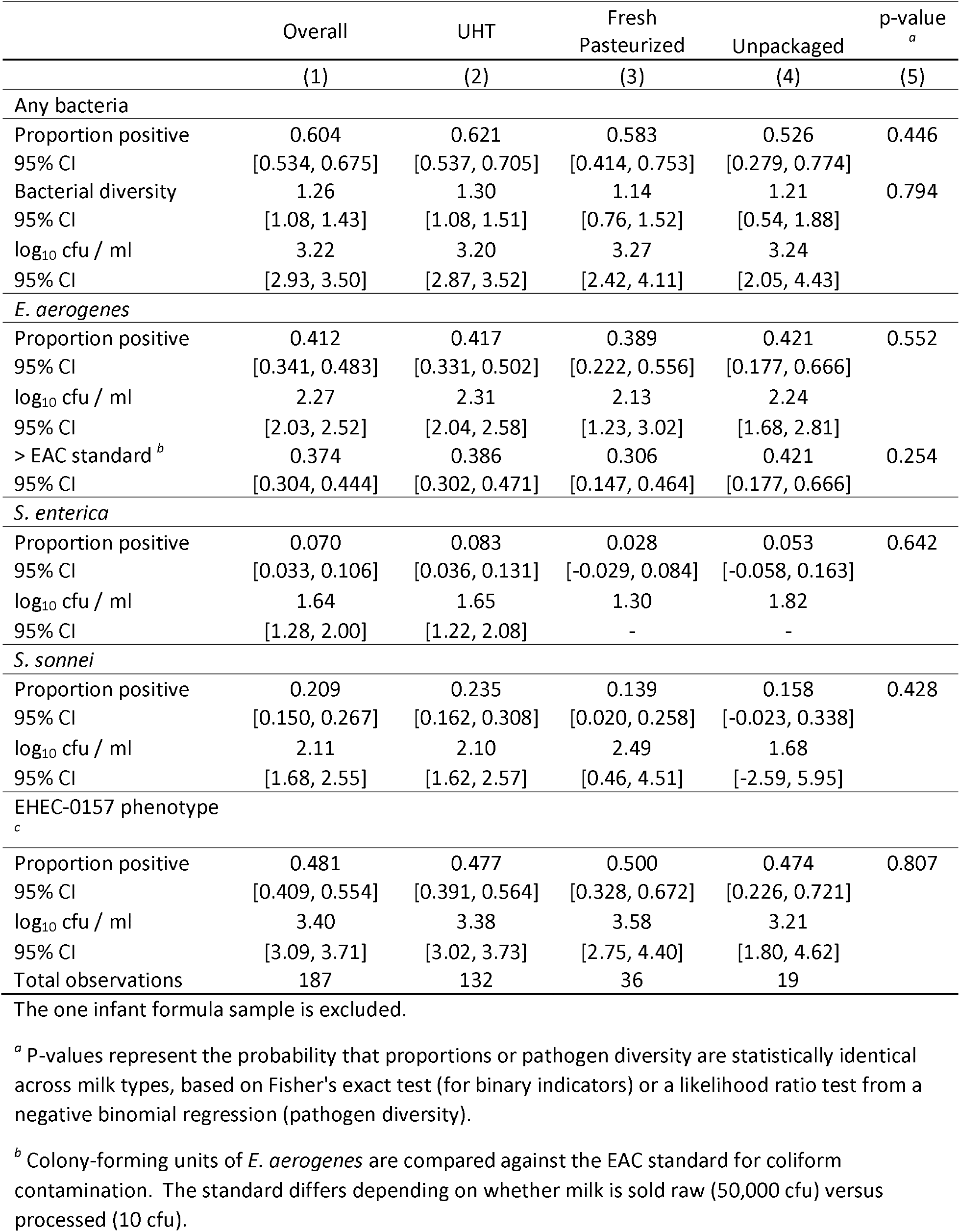
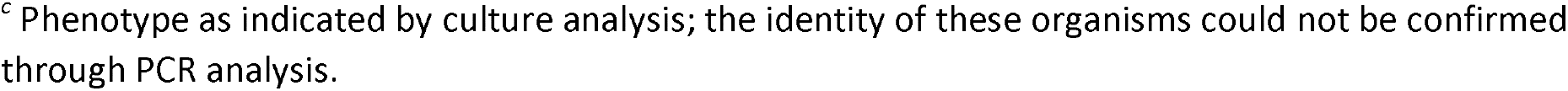
Microbial detection rates, diversity, and concentration in 187 infant food samples matched to fluid vendor milk samples, by milk treatment type.

|  | Overall | UHT | Fresh<br>Pasteurized | Unpackaged | p-value<sup>a</sup> |
| --- | --- | --- | --- | --- | --- |
|  | (1) | (2) | (3) | (4) | (5) |
| *Any bacteria* |  |  |  |  |  |
| Proportion positive | 0.604 | 0.621 | 0.583 | 0.526 | 0.446 |
| 95% CI | [0.534, 0.675] | [0.537, 0.705] | [0.414, 0.753] | [0.279, 0.774] |  |
| Bacterial diversity | 1.26 | 1.30 | 1.14 | 1.21 | 0.794 |
| 95% CI | [1.08, 1.43] | [1.08, 1.51] | [0.76, 1.52] | [0.54, 1.88] |  |
| log<sub>10</sub> cfu / ml | 3.22 | 3.20 | 3.27 | 3.24 |  |
| 95% CI | [2.93, 3.50] | [2.87, 3.52] | [2.42, 4.11] | [2.05, 4.43] |  |
| *E. aerogenes* |  |  |  |  |  |
| Proportion positive | 0.412 | 0.417 | 0.389 | 0.421 | 0.552 |
| 95% CI | [0.341, 0.483] | [0.331, 0.502] | [0.222, 0.556] | [0.177, 0.666] |  |
| log<sub>10</sub> cfu / ml | 2.27 | 2.31 | 2.13 | 2.24 |  |
| 95% CI | [2.03, 2.52] | [2.04, 2.58] | [1.23, 3.02] | [1.68, 2.81] |  |
| > EAC standard<sup>b</sup> | 0.374 | 0.386 | 0.306 | 0.421 | 0.254 |
| 95% CI | [0.304, 0.444] | [0.302, 0.471] | [0.147, 0.464] | [0.177, 0.666] |  |
| *S. enterica* |  |  |  |  |  |
| Proportion positive | 0.070 | 0.083 | 0.028 | 0.053 | 0.642 |
| 95% CI | [0.033, 0.106] | [0.036, 0.131] | [-0.029, 0.084] | [-0.058, 0.163] |  |
| log<sub>10</sub> cfu / ml | 1.64 | 1.65 | 1.30 | 1.82 |  |
| 95% CI | [1.28, 2.00] | [1.22, 2.08] | - | - |  |
| *S. sonnei* |  |  |  |  |  |
| Proportion positive | 0.209 | 0.235 | 0.139 | 0.158 | 0.428 |
| 95% CI | [0.150, 0.267] | [0.162, 0.308] | [0.020, 0.258] | [-0.023, 0.338] |  |
| log<sub>10</sub> cfu / ml | 2.11 | 2.10 | 2.49 | 1.68 |  |
| 95% CI | [1.68, 2.55] | [1.62, 2.57] | [0.46, 4.51] | [-2.59, 5.95] |  |
| EHEC-0157 phenotype<sup>c</sup> |  |  |  |  |  |
| Proportion positive | 0.481 | 0.477 | 0.500 | 0.474 | 0.807 |
| 95% CI | [0.409, 0.554] | [0.391, 0.564] | [0.328, 0.672] | [0.226, 0.721] |  |
| log<sub>10</sub> cfu / ml | 3.40 | 3.38 | 3.58 | 3.21 |  |
| 95% CI | [3.09, 3.71] | [3.02, 3.73] | [2.75, 4.40] | [1.80, 4.62] |  |
| Total observations | 187 | 132 | 36 | 19 |  |

The one infant formula sample is excluded.
<sup>a</sup> P-values represent the probability that proportions or pathogen diversity are statistically identical across milk types, based on Fisher's exact test (for binary indicators) or a likelihood ratio test from a negative binomial regression (pathogen diversity).
<sup>b</sup> Colony-forming units of *E. aerogenes* are compared against the EAC standard for coliform contamination. The standard differs depending on whether milk is sold raw (50,000 cfu) versus processed (10 cfu).
<sup>c</sup> Phenotype as indicated by culture analysis; the identity of these organisms could not be confirmed through PCR analysis.

Splitting the sample by milk type, rates of detection for all of the bacteria studied were higher in infant food prepared with UHT milk than in its vendor milk source (p=0.039 for 5. *enterica;* for all other organisms p<0.001). Infant food prepared with fresh pasteurized milk was also more likely to contain at least one type of bacteria than its vendor milk source (p=0.035), though only one organism, the EHEC-0157 phenotype, was significantly more likely to be detected in infant food than in the paired vendor source (p=0.023). In contrast, detection of any bacteria was less likely in infant food prepared with unpackaged milk compared to paired vendor source (p=0.004). The difference in probability of detection between unpackaged vendor milk and paired infant food samples is significant for 5. *enterica* (p=0.008), 5. *sonnei* (p=0.001), and the EHEC-0157 phenotype (p=0.008), but not for *E. aerogenes* (p=0.388).

After adjusting models for how milk was used in infant feeding, 5. *enterica* was less likely to be detected in cooked foods prepared with milk (tea, porridge) compared to uncooked milk (pure milk or milk in cold cereal), holding the influence of milk type (UHT, pasteurized, unpackaged) constant (Table S2). This adjustment accounts for mixed infant foods to potentially increase in contamination due to adding other contaminated food ingredients, or to decrease in contamination due to cooking. Results are similar to those shown in Table 3.

### Sources of pathogens in infant food

Among 188 matched vended milk-infant food sample pairs from Safe Start control group caregivers, *E. aerogenes* was detected in 56% of the 27 infant foods prepared with *E. aerogenes* positive vendor milk, 5. *enterica* was detected in 14% of the 14 infant foods prepared with 5. *enterica* positive vendor milk, 5. *sonnei* was detected in 25% of the 16 infant foods prepared with 5. *sonnei* positive vendor milk, and the EHEC-0157 phenotype was found in 61% of infant foods prepared with vendor milk prepared with EHEC-0157 phenotype positive vendor milk (Table 4).

**Table 4.**
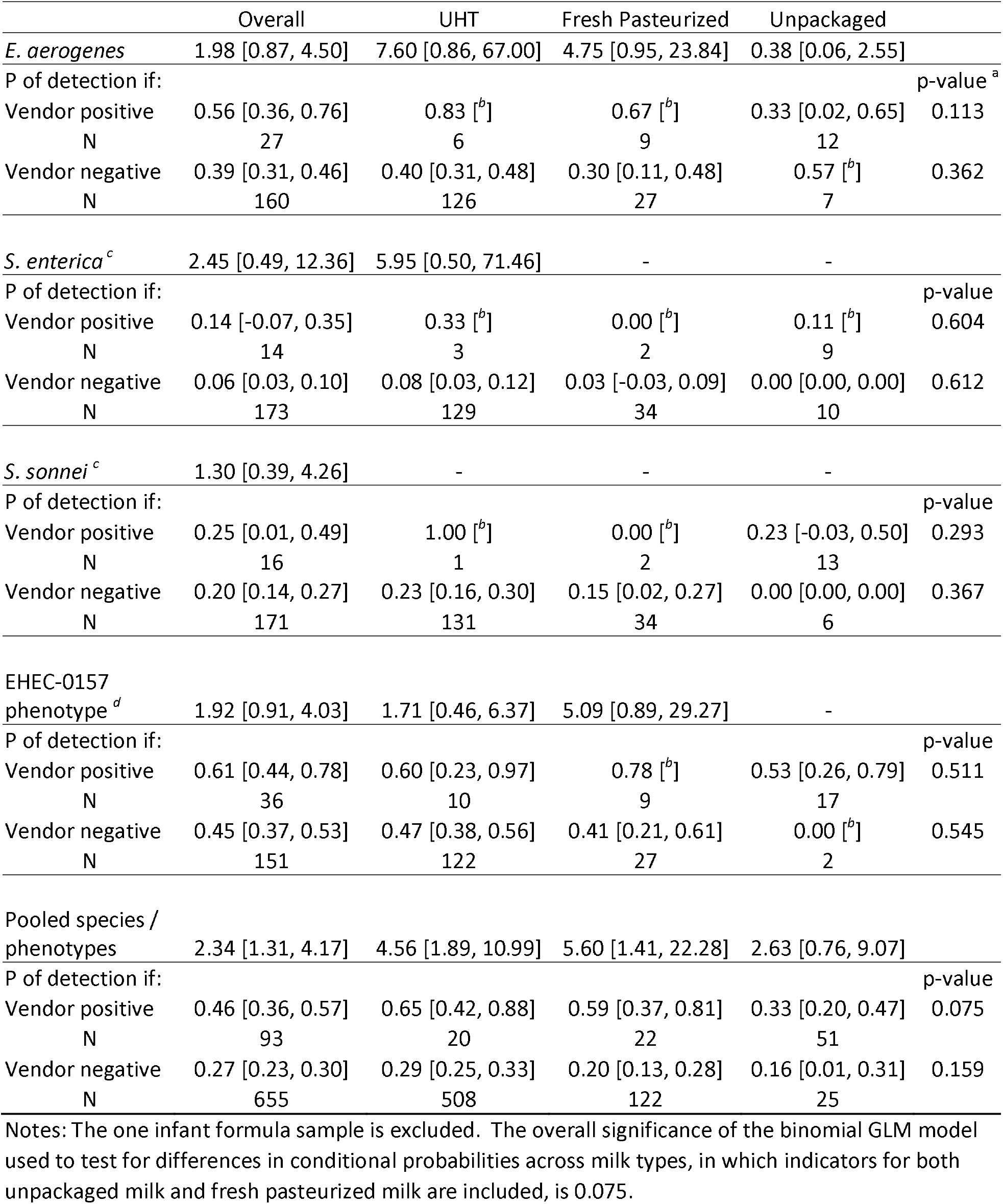
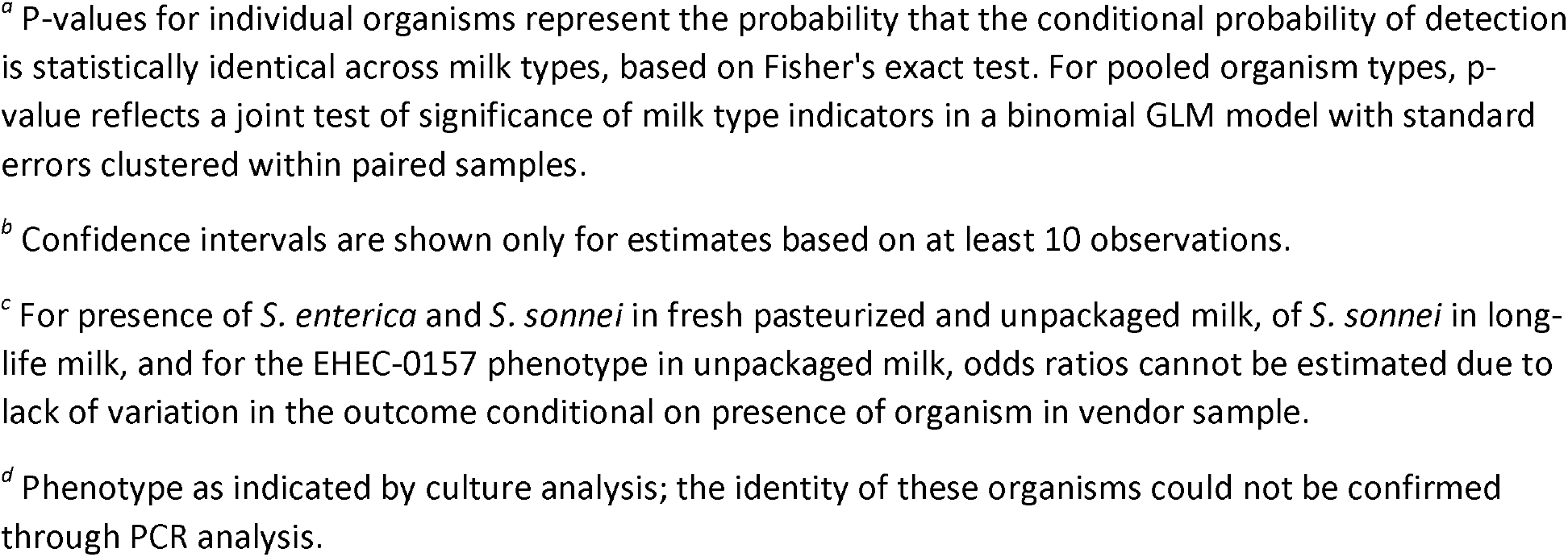
Odds ratio (95% Cl) of bacterial detection in infant food based upon vendor milk contamination status by type of milk product, and sub-group conditional probabilities (95% Cl) that organism is detected in infant food sample

|  | Overall | UHT | Fresh Pasteurized | Unpackaged |  |
| --- | --- | --- | --- | --- | --- |
| *E. aerogenes* | 1.98 [0.87, 4.50] | 7.60 [0.86, 67.00] | 4.75 [0.95, 23.84] | 0.38 [0.06, 2.55] |  |
| P of detection if: |  |  |  |  | p-value <sup>a</sup> |
| Vendor positive | 0.56 [0.36, 0.76] | 0.83 [<sup>b</sup>] | 0.67 [<sup>b</sup>] | 0.33 [0.02, 0.65] | 0.113 |
| N | 27 | 6 | 9 | 12 |  |
| Vendor negative | 0.39 [0.31, 0.46] | 0.40 [0.31, 0.48] | 0.30 [0.11, 0.48] | 0.57 [<sup>b</sup>] | 0.362 |
| N | 160 | 126 | 27 | 7 |  |
| *S. enterica* <sup>c</sup> | 2.45 [0.49, 12.36] | 5.95 [0.50, 71.46] | - | - |  |
| P of detection if: |  |  |  |  | p-value |
| Vendor positive | 0.14 [-0.07, 0.35] | 0.33 [<sup>b</sup>] | 0.00 [<sup>b</sup>] | 0.11 [<sup>b</sup>] | 0.604 |
| N | 14 | 3 | 2 | 9 |  |
| Vendor negative | 0.06 [0.03, 0.10] | 0.08 [0.03, 0.12] | 0.03 [-0.03, 0.09] | 0.00 [0.00, 0.00] | 0.612 |
| N | 173 | 129 | 34 | 10 |  |
| *S. sonnei* <sup>c</sup> | 1.30 [0.39, 4.26] | - | - | - |  |
| P of detection if: |  |  |  |  | p-value |
| Vendor positive | 0.25 [0.01, 0.49] | 1.00 [<sup>b</sup>] | 0.00 [<sup>b</sup>] | 0.23 [-0.03, 0.50] | 0.293 |
| N | 16 | 1 | 2 | 13 |  |
| Vendor negative | 0.20 [0.14, 0.27] | 0.23 [0.16, 0.30] | 0.15 [0.02, 0.27] | 0.00 [0.00, 0.00] | 0.367 |
| N | 171 | 131 | 34 | 6 |  |
| EHEC-0157<br>phenotype <sup>d</sup> | 1.92 [0.91, 4.03] | 1.71 [0.46, 6.37] | 5.09 [0.89, 29.27] | - |  |
| P of detection if: |  |  |  |  | p-value |
| Vendor positive | 0.61 [0.44, 0.78] | 0.60 [0.23, 0.97] | 0.78 [<sup>b</sup>] | 0.53 [0.26, 0.79] | 0.511 |
| N | 36 | 10 | 9 | 17 |  |
| Vendor negative | 0.45 [0.37, 0.53] | 0.47 [0.38, 0.56] | 0.41 [0.21, 0.61] | 0.00 [<sup>b</sup>] | 0.545 |
| N | 151 | 122 | 27 | 2 |  |
| Pooled species /<br>phenotypes | 2.34 [1.31, 4.17] | 4.56 [1.89, 10.99] | 5.60 [1.41, 22.28] | 2.63 [0.76, 9.07] |  |
| P of detection if: |  |  |  |  | p-value |
| Vendor positive | 0.46 [0.36, 0.57] | 0.65 [0.42, 0.88] | 0.59 [0.37, 0.81] | 0.33 [0.20, 0.47] | 0.075 |
| N | 93 | 20 | 22 | 51 |  |
| Vendor negative | 0.27 [0.23, 0.30] | 0.29 [0.25, 0.33] | 0.20 [0.13, 0.28] | 0.16 [0.01, 0.31] | 0.159 |
| N | 655 | 508 | 122 | 25 |  |

Notes: The one infant formula sample is excluded. The overall significance of the binomial GLM model used to test for differences in conditional probabilities across milk types, in which indicators for both unpackaged milk and fresh pasteurized milk are included, is 0.075.
<sup>a</sup> P-values for individual organisms represent the probability that the conditional probability of detection is statistically identical across milk types, based on Fisher's exact test. For pooled organism types, p-value reflects a joint test of significance of milk type indicators in a binomial GLM model with standard errors clustered within paired samples.
<sup>b</sup> Confidence intervals are shown only for estimates based on at least 10 observations.
<sup>c</sup> For presence of *S. enterica* and *S. sonnei* in fresh pasteurized and unpackaged milk, of *S. sonnei* in long-life milk, and for the EHEC-0157 phenotype in unpackaged milk, odds ratios cannot be estimated due to lack of variation in the outcome conditional on presence of organism in vendor sample.
<sup>d</sup> Phenotype as indicated by culture analysis; the identity of these organisms could not be confirmed through PCR analysis.

Odds ratios of detection based on the pooled sample for all milk types indicate that for any given bacterial species or phenotype, its presence in vendor milk does not significantly predict its presence in infant food prepared with that milk. The low frequency of bacterial detection in vendor samples limits statistical power to detect differences in the probability of contamination of infant food conditional on vendor milk status by organism.

Pooling the microbial indicators such that observations are at the sample-organism level and allowing the detection of different bacterial species or phenotypes to be correlated within paired samples, yielded 748 observations in total, 4 per paired sample of vendor milk and infant food (Table 4, final panel). With this larger number of positive vendor samples (n = 93), we find that the presence of a particular bacterial species or phenotype in any type of vendor milk is significantly associated with greater risk of contamination with that same species in infant food (OR = 2.34, 95% CI: 1.31, 4.17). This is the case for UHT (OR = 4.56, 95% CI 1.89, 10.99) and fresh pasteurized milk (OR = 5.60, 95% CI: 1.41, 22.3), but not for unpackaged milk (OR = 2.63, 95% CI: 0.76, 9.07). The probability of a bacterial species co-detection in infant food, conditional on detection in the vended source, is significantly lower (p=0.029; not shown) in unpackaged milk, at 0.33 (95% CI: 0.20, 0.47), than in UHT milk, at 0.65 (95% CI: 0.42, 0.88).

The change in bacteria-specific contamination concentrations between vendor and infant food samples by type of milk was examined to further understand the nature of household handling on bacterial transmission from vendor milk. A large share of observations was negative for each species of bacteria in both the vendor milk and infant food sample. The distribution of changes in concentration was thus not amenable to analysis as a continuous variable. We therefore discretize the change in concentration to a four-value categorical variable: not detected in either sample, lower concentration in infant food than vendor milk, higher concentration in infant food than vendor milk, and maximum limit of detection in both samples. In only three cases were both the matched vendor milk and infant food sample contaminated at the maximum limit of detection. In all three cases, this occurred for the EHEC-0157 phenotype; one sample was UHT milk and two were unpackaged milk.

Table 5 shows the proportion of 187 matched samples by fluid milk type and bacterial species or phenotypes, in which microbial concentration was consistently zero, higher in the vended milk sample, and higher in the infant food sample. For all four types of bacteria, the probability that both samples test negative is lowest in unpackaged milk. Among matched samples for which the level of concentration differs between vended milk and infant food, food prepared with UHT milk is consistently more contaminated than the milk with which it was prepared. Infant food prepared with raw milk, in contrast, is consistently less contaminated by the time it is consumed by an infant, relative to the milk at purchase. Fresh pasteurized milk generally follows the same pattern as UHT milk, with the exception of 5. *enterica*, for which microbial concentration is higher in infant food than vendor milk.

**Table 5.**
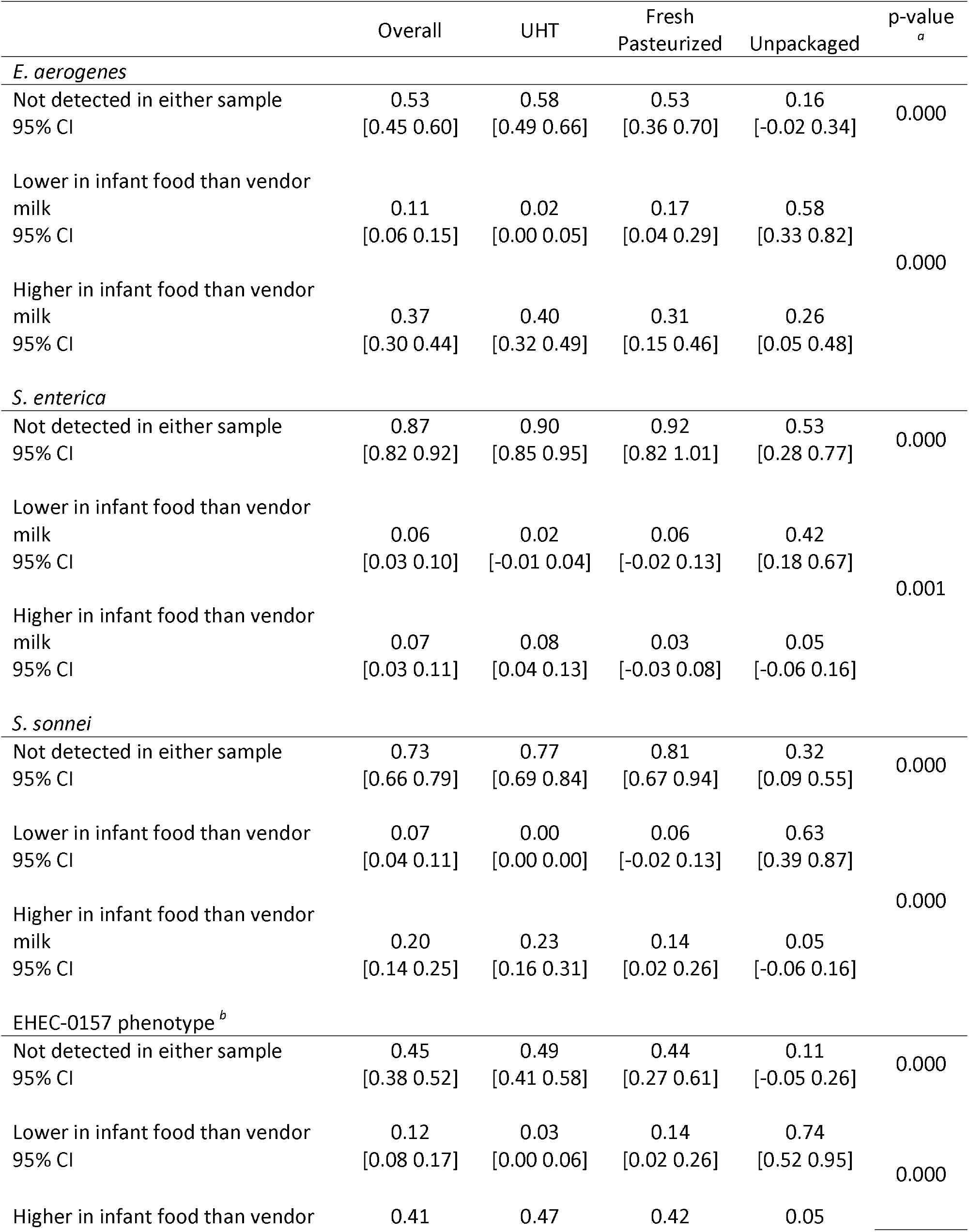
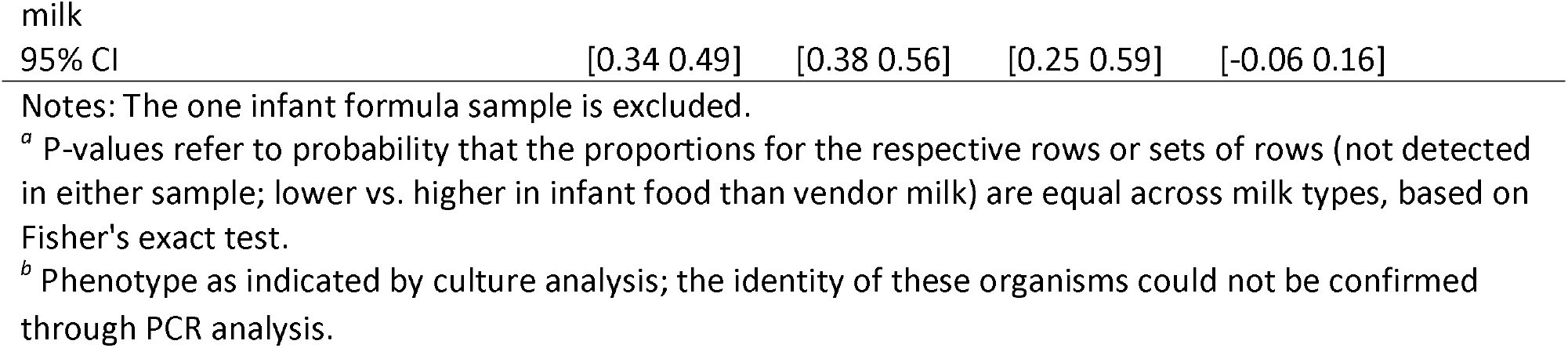
Proportion of samples by categorical change in microbial concentration between fluid vendor milk and infant samples for 187 matched samples.

|  | Overall | UHT | Fresh<br>Pasteurized | Unpackaged | p-value<sup>a</sup> |
| --- | --- | --- | --- | --- | --- |
| *E. aerogenes* |  |  |  |  |  |
| Not detected in either sample<br>95% CI | 0.53<br>[0.45 0.60] | 0.58<br>[0.49 0.66] | 0.53<br>[0.36 0.70] | 0.16<br>[-0.02 0.34] | 0.000 |
| Lower in infant food than vendor<br>milk<br>95% CI | 0.11<br>[0.06 0.15] | 0.02<br>[0.00 0.05] | 0.17<br>[0.04 0.29] | 0.58<br>[0.33 0.82] | 0.000 |
| Higher in infant food than vendor<br>milk<br>95% CI | 0.37<br>[0.30 0.44] | 0.40<br>[0.32 0.49] | 0.31<br>[0.15 0.46] | 0.26<br>[0.05 0.48] |  |
| *S. enterica* |  |  |  |  |  |
| Not detected in either sample<br>95% CI | 0.87<br>[0.82 0.92] | 0.90<br>[0.85 0.95] | 0.92<br>[0.82 1.01] | 0.53<br>[0.28 0.77] | 0.000 |
| Lower in infant food than vendor<br>milk<br>95% CI | 0.06<br>[0.03 0.10] | 0.02<br>[-0.01 0.04] | 0.06<br>[-0.02 0.13] | 0.42<br>[0.18 0.67] | 0.001 |
| Higher in infant food than vendor<br>milk<br>95% CI | 0.07<br>[0.03 0.11] | 0.08<br>[0.04 0.13] | 0.03<br>[-0.03 0.08] | 0.05<br>[-0.06 0.16] |  |
| *S. sonnei* |  |  |  |  |  |
| Not detected in either sample<br>95% CI | 0.73<br>[0.66 0.79] | 0.77<br>[0.69 0.84] | 0.81<br>[0.67 0.94] | 0.32<br>[0.09 0.55] | 0.000 |
| Lower in infant food than vendor<br>95% CI | 0.07<br>[0.04 0.11] | 0.00<br>[0.00 0.00] | 0.06<br>[-0.02 0.13] | 0.63<br>[0.39 0.87] | 0.000 |
| Higher in infant food than vendor<br>milk<br>95% CI | 0.20<br>[0.14 0.25] | 0.23<br>[0.16 0.31] | 0.14<br>[0.02 0.26] | 0.05<br>[-0.06 0.16] |  |
| EHEC-0157 phenotype<sup>b</sup> |  |  |  |  |  |
| Not detected in either sample<br>95% CI | 0.45<br>[0.38 0.52] | 0.49<br>[0.41 0.58] | 0.44<br>[0.27 0.61] | 0.11<br>[-0.05 0.26] | 0.000 |
| Lower in infant food than vendor<br>95% CI | 0.12<br>[0.08 0.17] | 0.03<br>[0.00 0.06] | 0.14<br>[0.02 0.26] | 0.74<br>[0.52 0.95] | 0.000 |
| Higher in infant food than vendor | 0.41 | 0.47 | 0.42 | 0.05 |  |

|  |  |  |  |  |
| --- | --- | --- | --- | --- |
| milk |  |  |  |  |
| 95% CI | [0.34 0.49] | [0.38 0.56] | [0.25 0.59] | [-0.06 0.16] |

Notes: The one infant formula sample is excluded.
<sup>a</sup> P-values refer to probability that the proportions for the respective rows or sets of rows (not detected in either sample; lower vs. higher in infant food than vendor milk) are equal across milk types, based on Fisher's exact test.
<sup>b</sup> Phenotype as indicated by culture analysis; the identity of these organisms could not be confirmed through PCR analysis.

## DISCUSSION

This study compared bacterial contamination patterns in infant foods and their vended cow’s milk sources to assess the role of food system versus household contamination sources in infant food. We draw several notable conclusions from this study. First, as generally expected, consumption of pasteurized milk in this setting in Kenya was found to be microbiologically safer than unpacked or raw milk. However, there are problems with the dairy system. It is noteworthy that a majority of these urban low-income caregivers choose to purchase long-life milk, the most expensive type available, for their infants. We do not have data on milk consumed by other family members, and it is possible that the apparent consumer preference for pasteurized and UHT milk is specific to infant feeding. Representative data on milk consumption patterns in peri-urban Kisumu indicate that 60% of households regularly consume processed milk, but do not specify which household members consume this milk, or distinguish between UHT and fresh pasteurized milk ^21^. However, of the 396 samples of milk obtained by caregivers of infants in Kisumu, 8.6% (34 samples) were found to be contaminated with *Shigella sonnei, Salmonella enterica*, or both pathogens. While unpackaged milk samples were far more likely to be contaminated with one of the pathogens, pathogen contamination in UHT milk (2.2%) and fresh pasteurized milk (4.8%) were also nontrivial. Second, any benefits from purchasing UHT and fresh pasteurized disappear by the time infants are fed. The frequency of bacterial contamination in food was similar for all milk types.

Examination of co-detection patterns in matched vendor milk-infant food pairs across multiple individual bacterial species demonstrated that vendor milk safety issues and household hygiene both contributed to this trend. Microbial contamination of infant food most commonly arose during handling of food within the household, or from other food ingredients to which milk was added. This reinforces the importance of household-based interventions that can improve infant food preparation, feeding, and storage hygiene conditions. Yet, in multiple cases 5. *sonnei* and 5. *enterica* were detected in matched vended milk sources and infant foods. Pooling pathogenic and indicator organisms, the proportion of cases in which an organism present in vendor milk was also present after household handling and storage was highest in UHT milk, similar in fresh pasteurized milk, and lowest in unpackaged milk. Changes in microbial concentration between milk purchase and collection of infant food samples showed the same pattern, with infant food prepared with UHT milk consistently more contaminated than the milk with which it was prepared, and infant food prepared with unpackaged milk consistently less contaminated (if any change was observed). These patterns do suggest that how caregivers handle milk differs based on how it is processed and whether it is packaged or unpackaged ^8^.

The significance of the pathogens detected in food through this study in self-reported diarrhea and infection outcomes observed in the Safe Start trial remains to be assessed. Foodborne disease accounts for a significant amount of diarrheal illness in children in Kenya ^22^. Data compiled by the Foodborne Disease Burden Epidemiology Reference Group of the WHO suggests that 5. *enterica* and *Shigella* spp. are among the first and sixth most common causes of diarrheal foodborne illness ^1 22^. While challenge studies among healthy adult volunteers in low burden, high-income countries report infectious doses of 10^5^-10^10^ organisms, data from outbreaks in similar settings indicate that the infective dose of 5. *enterica* may be as low as 100 organisms, likely due to higher susceptibility among vulnerable groups such as infants, the elderly, and the sick ^23^. The concentrations of 5. *enterica* observed in positive vendor samples in this study would be capable of causing disease if infants consume at least 25 mL of milk. Between 10-200 5. *sonnei* organisms can cause disease in healthy adults ^24^. Minimum thresholds for infection would have been met by consuming 25 mL of milk for 92% of the 23 samples in which 5. *sonnei* was detected. For infants, even samples with the lowest level of contamination could be dangerous. The findings thus indicate cause for concern about the safety of both raw and pasteurized sold milk in Kisumu for infants.

A limitation of this study is our analysis of pathogen transmission from purchased milk to infant food was limited by relatively low rates of detection of pathogens in vended milk sources. Additionally, we used cost-effective phenotype or gene-level typing methods to assess probable relatedness between vendor milk and infant food matched samples. Determination of base pair relatedness via sequencing would have improved certainty in transmission conclusions but was beyond the scope of this project. The strength of our conclusions instead relies upon observing repeat patterns for a variety of bacteria species including common indicators and relatively rare pathogens. A shocking number of infant foods were contaminated by a bacteria phenotype matching EHEC 0157 (sorbitol negative, β-D-glucuronidase negative). EHEC 0157 identity was not validated by PCR screening and remains unidentified.

## CONCLUSION

The presence of bacterial pathogens in the Kenyan milk supply suggests dairy food systems in Kenya could contribute to pediatric foodborne outbreaks, especially if those sources are not re-boiled by consumers. Differing rates of compliance with the EAC coliform standard for processed milk across brands of UHT milk indicates that actions by milk processing firms do affect levels of microbial contamination. Contamination of widely distributed packaged foods with pathogens indicates the potential for exposing a large share of the population. However, rates of pathogen detection at point of purchase were relatively low compared to household contamination frequencies, implying an opportunity for larger health gains by improving food hygiene at the household level. Joint interventions addressing food safety in both the milk supply chain and among consumers could reduce the risk of infant exposure to foodborne pathogens.

## Data Availability

All data referred to in the manuscript on food sources used to make infant food will be made fully available by request. Any data referred to as household infant food data will be made available after publication of the primary results of the Safe Start study.

## ACKNOWLEDGMENTS

This study was supported by the CGIAR Research Program on Agriculture for Nutrition and Health (A4NH), hosted by IFPRI, and the Dutch Ministry of Foreign Affairs through SNV (Netherlands Development Organization) under the Voices for Change Partnership. The Safe Start trial, on which the study built, was funded by the United Kingdom Department for International Development (DFID) through the SHARE Research Consortium. We appreciate the support of Julius Otieno, Edwin Attitwa, Bonphace Okoth, John Agira, and Horace Omondi who assisted with the collection of data, implementation of microbial assays, and data management. All authors declare that they have no competing financial interests in the publication of this study.

## Author Contributions

SW, NJ, DW and SS performed the research. SW, NJ, HH and TL designed the research study. HH and SS contributed essential reagents or tools. VH and DW analysed the data. VH and KKB wrote the paper

